# Improving Diagnostic Accuracy of Routine EEG for Epilepsy using Deep Learning

**DOI:** 10.1101/2025.01.13.25320425

**Authors:** Émile Lemoine, Denahin Toffa, An Qi Xu, Jean-Daniel Tessier, Mezen Jemel, Frédéric Lesage, Dang K. Nguyen, Elie Bou Assi

**Author notes:** **Corresponding author:** Elie Bou Assi, 1051 rue Sanguinet, Montréal, Québec. H2X 3E4. These authors should be considered co-senior authors (Dang K. Nguyen, Elie Bou Assi).

## Abstract

**Background and Objectives:** The diagnostic yield of routine EEG in epilepsy is limited by low sensitivity and the potential for misinterpretation of interictal epileptiform discharges (IEDs). Our objective is to develop, train, and validate a deep learning model that can identify epilepsy from routine EEG recordings, complementing traditional IED-based interpretation.

**Methods:** This is a retrospective cohort study of diagnostic accuracy. All consecutive patients undergoing routine EEG at our tertiary care center between January 2018 and September 2019 were included. EEGs recorded between July 2019 and September 2019 constituted a temporally shifted testing cohort. The diagnosis of epilepsy was established by the treating neurologist at the end of the available follow-up period, based on clinical file review. Original EEG reports were reviewed for IEDs. We developed seven novel deep learning models based on Vision Transformers (ViT) and Convolutional Neural Networks (CNN), training them to classify raw EEG recordings. We compared their performance to IED-based interpretation and two previously proposed machine learning methods.

**Results:** The study included 948 EEGs from 846 patients (820 EEGs/728 patients in training/validation, 128 EEGs/118 patients in testing). Median follow-up was 2.2 years and 1.7 years in each cohort, respectively. Our flagship ViT model, DeepEpilepsy, achieved an area under the receiver operating characteristic curve (AUROC) of 0.76 (95% CI: 0.69–0.83), outperforming IED-based interpretation (0.69; 0.64–0.73) and previous methods. Combining DeepEpilepsy with IEDs increased the AUROC to 0.83 (0.77–0.89).

**Discussion:** DeepEpilepsy can identify epilepsy on routine EEG independently of IEDs, suggesting that deep learning can detect novel EEG patterns relevant to epilepsy diagnosis. Further research is needed to understand the exact nature of these patterns and evaluate the clinical impact of this increased diagnostic yield in specific settings.

## Introduction

The diagnosis of epilepsy is notoriously challenging. It relies on the occurrence of either two seizures more than 24h apart, one seizure and a high risk of another, or the presence of an epilepsy syndrome.^1^ Despite this clear definition, the rate of misdiagnosis remains high.^2,3^ A key issue is the lack of robust and validated interictal biomarkers,^4–6^ making the diagnosis highly dependent on the ability to collect a clear clinical history and accurately interpret the electroencephalogram (EEG).

The EEG can capture ictal and interictal activity that is highly specific for epilepsy. It is cost-effective and technically straightforward, with standard acquisition protocols that have been put in place by the International League Against Epilepsy.^7,8^ However, its diagnostic yield is hampered by low sensitivity^9^ and only moderate interrater reliability.^10^ Consequently, the EEG has limitations as a diagnostic tool in patients with suspected seizures, with EEG misinterpretation contributing to diagnostic errors in epilepsy.^11^

In recent decades, efforts have focused on overcoming the limitations of traditional EEG interpretation by identifying alternative epilepsy biomarkers within the EEG through computational methods.^12–16^ More recently, Deep learning (DL) has since revolutionized the analysis of complex signals. DL models can autonomously extract features from time-series or images by optimizing millions of parameters on large datasets. DL has been applied to EEG to decode brain signals for brain-computer interface,^17^ predict delirium,^18^ and automatically detect IEDs.^19,20^ Despite these advancements, studies that attempt to detect epilepsy on EEG remain disconnected from clinical practice, often using unrepresentative samples or lacking robust validation.^13^ As a result, the true diagnostic accuracy of these approaches is uncertain, and clinical translation is still awaited.

Our group recently demonstrated that machine learning could predict the risk of seizures in the year following a routine EEG.^21^ This method could also predict the diagnosis of epilepsy from the EEG alone with an area under the receiver operating characteristic curve (AUROC) of 0.63. The extraction of pre-defined features has limited capacity to capture the complex brain dynamics underlying epilepsy, leading us to hypothesize that DL could substantially enhance these performances.

The present study builds on these findings and seeks to address these questions: can modern DL models detect epilepsy on interictal EEG, even in the absence of IEDs? What are the potential diagnostic performances of a DL-assisted EEG interpretation for epilepsy? And what sample size is required to train such models?

## Methods

### Study design

This is a retrospective study on a consecutive cohort of patients undergoing routine EEG in a single tertiary care center in Montreal, Canada.

### Participants

We included all patients who underwent a routine EEG (20- to 60-minute, with or without sleep deprivation) between January 2018 and September 2019 at the Centre Hospitalier de l’Université de Montréal (CHUM). Exclusion criteria were the absence of follow-up after the EEG, an uncertain diagnosis of epilepsy at the end of the available follow-up period, or an EEG performed in a hospitalized patient. Under a prespecified protocol, one neurology resident (EL) and three students (AQ, MJ, JDT) collected data from the electronic health record for each visit, including baseline characteristics (age, sex), co-morbidities, number of antiseizure medications, and presence of a focal lesion on neuroimaging. They also reviewed the EEG report for the presence of IED(s) and abnormal background slowing. All clinical information was stored on a REDCap database hosted on the CHUM research center’s servers.

We separated the cohort into two independent subsets according to the date of the EEG. Recordings before July 15, 2019, comprised the training and validation set, while recordings after July 15, 2019, comprised the testing set. We excluded from the testing set any recording from a patient already included in the training and validation set. The training and validation set was further separated into a training set and a validation set in a random fashion (80%/20% split).

### Test Methods

#### Reference Standard

The reference standard is the diagnosis of epilepsy according to the treating physician at the end of the available follow-up period. This diagnosis is based on the ILAE definition of epilepsy, i.e. having had two unprovoked seizures more than 24h apart or one unprovoked seizure and be considered at high (> 60%) risk of seizure recurrence, or being diagnosed with an epilepsy syndrome.^1^ The final diagnosis at the end of the follow up period was used, as opposed to the speculated diagnosis at the time of the EEG, because the follow up period provides additional information such as imaging, additional EEG recordings, video-monitoring admissions, and seizure recurrences.

#### EEG recording

EEGs were recorded using a standardized protocol on a Nihon Kohden EEG system, following national recommendations.^22^ Awake EEGs, 20–30 minutes long, were recorded at 200 Hz with 19 electrodes arranged with the 10-20 system. They included two 90-second periods of hyperventilation (except in patients > 80 years old, uncooperative, or with medical contraindications) and photic stimulation from 4 Hz to 22 Hz. Patients were also instructed to open or close their eyes at several times. Sleep deprived recordings lasted 60 minutes, with the same activation procedures. Technologists annotated the EEG in real-time. For this study, EEGs were converted to an average referential montage (A1-A2), saved to EDF format, and stored on the CHUM research center’s server for analysis.

#### Automated processing of EEG and classification

The index test is the classification of the EEG recordings using machine learning. We developed DeepEpilepsy, a Vision Transformer (ViT) model that takes raw EEG segments as input and outputs a probability of the diagnosis of epilepsy (**Figure 1**). EEGs were segmented into overlapping 10- or 30- second windows and directly used as input into the DL models. To enhance model generalization, we applied a random data augmentation algorithm during training.^4^ For each segment, an augmentation was drawn randomly from a set of transformations, which included filtering (band-pass, low-pass, high-pass), masking (channel, time), and adding noise (**eFigure 1**). These were applied with a 50% probability and randomized intensity. We performed a Bayesian hyperparameter search on the training and validation set to choose DeepEpilepsy’s final configuration. We also investigated different learning rates, weight decay, and batch size values. The final models were trained on the entire training and validation set. The optimization hyperparameters and model specifications are described in **eTable 4**.

**Figure 1:**
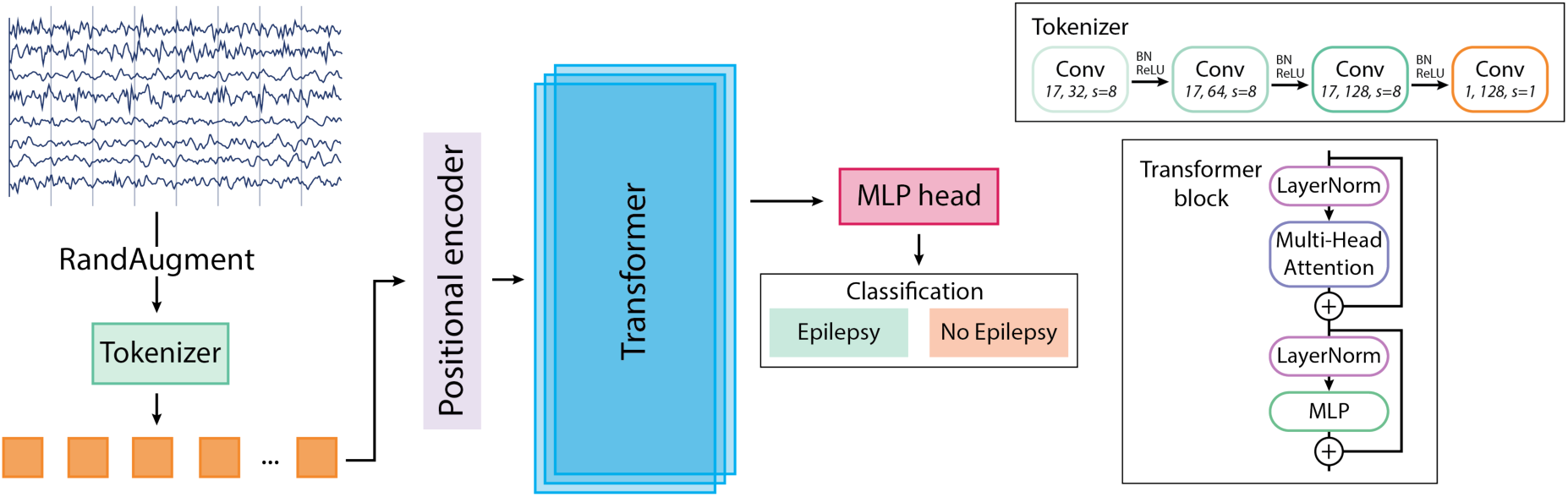
Details of the DeepEpilepsy Transformer model. The EEG is first processed through the RandAugm algorithm with 50% probability. A tokenizer is used (upper right: convolutional tokenizer) before positional encoding. The tokens are then input into a Transformer model. A MLP head classifies the embeddings from the Transformer according to the diagnosis of epilepsy. BN: Batch normalization; MLP: Multilayer perceptron; ReLU: Rectified linear unit.

In addition, we implemented other Deep Learning models (ViT and ConvNeXt), as well as two previously described methods: the ShallowConvNet inspired by the *Filter Bank Common Spatial Patterns* algorithm,^23^ and a feature-extraction framework relying on the extraction of linear and nonlinear EEG markers that are used as input into a classifier (LightGBM).^21^ These methods are described in details in **eMethods 1**.

To obtain the diagnostic performances, the final models/procedures were applied to the testing set. This resulted in a single predicted probability for each EEG segments. To obtain one prediction per EEG recording, we aggregated the predicted probabilities at the EEG-level using the median of the predicted values.

We further evaluated DeepEpilepsy in a specific subgroup of patients which were not yet diagnosed with epilepsy at the time of the index EEG (i.e., undergoing evaluation for suspected seizures). We also measured the performance bias across different subgroups: age groups (18–40, 40–60, and >60 years old), sex, presence of focal lesion, presence of IED (absence, presence, and uncertain), presence of slowing, and number of ASM (0, 1, ≥2).

### Analysis

We calculated the AUROC using the probabilistic predictions for each model, with 95% confidence intervals estimated using DeLong’s method (single prediction by patient).^24^ For comparison, we tested the classification performance of IEDs alone (presence vs. absence). We also tested a two-step classification using IEDs first (traditional EEG interpretation), followed by DeepEpilepsy if IEDs were absent (DL interpretation).

The optimal classification threshold was obtained using the validation cohort, minimizing the distance between the curve and the upper left corner of the ROC graph. This threshold was then applied to compute sensitivity, specificity, negative predictive value, and positive predictive value on the testing set.

We performed an exploratory analysis of the embeddings learned by DeepEpilepsy and ShallowConvNet to better understand the patterns captured by both models (**eMethods 2**).

### Sample size

Using Obuchowski’s method,^25^ with a 60% epilepsy prevalence, a power of 0.9, and a significance level of 0.0071 (adjusted from 0.05 divided by 7 DL models), a minimum of 126 EEGs is required to detect an AUROC of 0.70.

### Standard Protocol Approvals, Registrations, and Patient Consents

Ethics approval was granted by the CHUM Research Centre’s Research Ethics Board (REB) (Montreal, Canada, project number: 19.334). The REB waived informed consent due to the lack of diagnostic/therapeutic intervention and minimal risk to participants. All methods followed Canada’s Tri-Council Policy statement on Ethical Conduct for Research Involving Humans.

### Code and Data Availability

The code for the study will be available upon publication at the following address: https://gitlab.com/chum-epilepsy/dl_epilepsy_reeg. Anonymized data will be made available to qualified investigators upon reasonable request, conditional to the approval by our REB. The STARD checklist is provided as Supplementary material.

## Results

### Participants

After exclusion, 948 EEGs from 846 patients were included: 820 EEGs in the training/validation set (728 patients) and 128 EEGs in the testing set (118 patients), with no patient overlap. Before exclusion, 1,185 EEGs from 1 067 patients and 161 EEGs from 149 patients met the inclusion criteria for the training and testing cohorts, respectively. Reasons for exclusion were absence of follow-up after the EEG, uncertain diagnosis at the end of available follow-up, seizure during the EEG, and wrong EEG type (i.e., performed in a hospitalized patient) (**Figure 2**). Median age were 49 and 51.5 (IQR: 32–62 and 30–62.5) and the proportion of women were 51% and 62.5% in the training and testing cohorts, respectively. Median follow-up was 2.2 years (IQR: 1.0–2.9) and 1.7 years (IQR: 0.9–2.3). Epilepsy prevalence was 63% in both sets.

**Figure 2:**
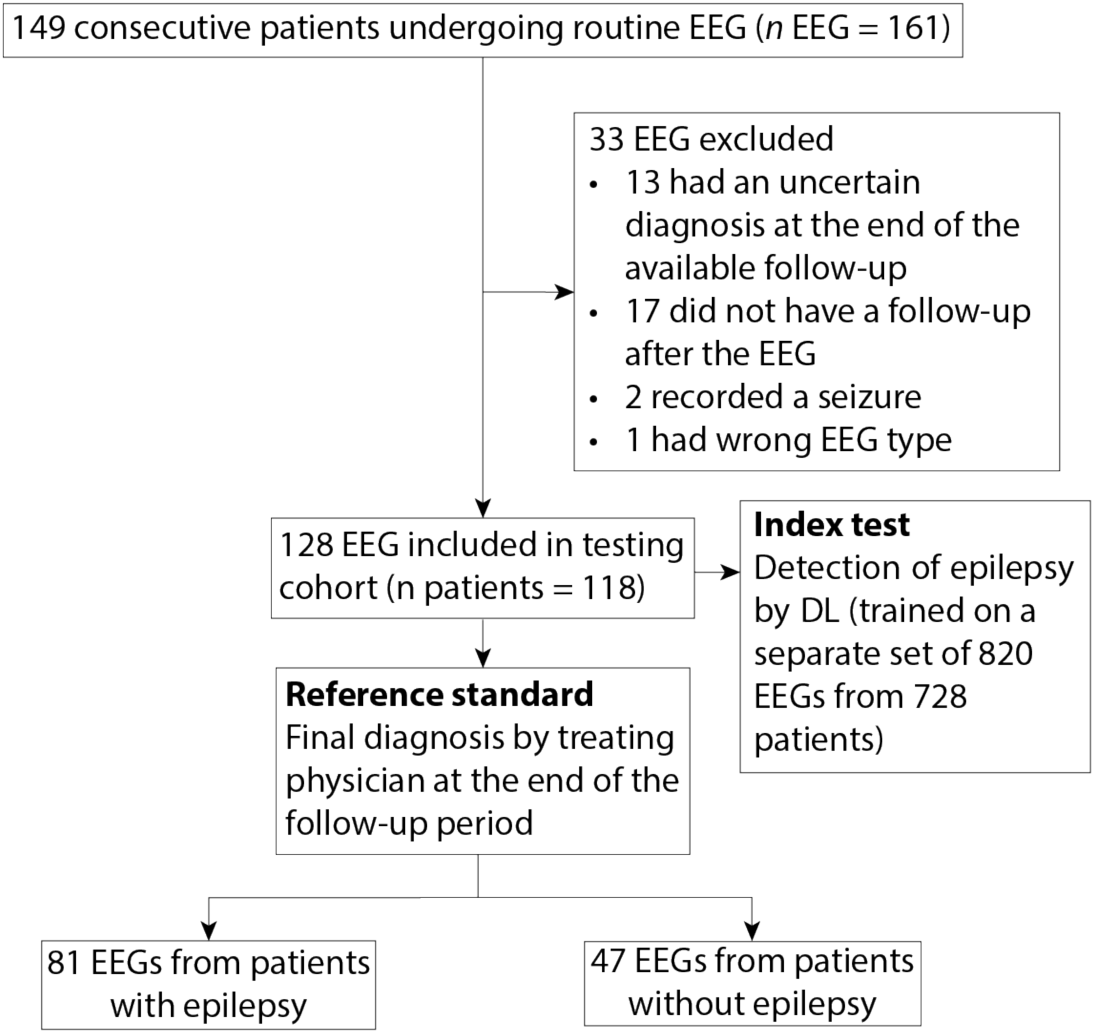
Flowchart of patients included in the testing cohort.

In the testing cohort, 75 patients (64%) had an uncertain diagnosis at the time of the EEG, 28 of which were eventually diagnosed with epilepsy. In the 47 others, the most common final diagnoses were syncope/faintness (11), dementia-related fluctuations (6), and non-specific sensitive symptoms (5). In EEGs from patients finally diagnosed with epilepsy, 10 showed IEDs and 6 had uncertain sharp transients (vs. 1 in patients without epilepsy). This subgroup is detailed in **eTable 5**.

### Test Results

The AUROC for the diagnosis of epilepsy in the testing cohort for every approach is pictured in **Figure 3**. For DeepEpilepsy, the AUROC was 0.76 (95%CI: 0.69–0.83). Using the threshold computed on the validation cohort (0.86), there were 75 true positives, 38 true negatives, 13 false positive, and 41 false negatives, equating to a sensitivity of 64.7%, a specificity of 74.5%, a positive predicted value (PPV) of 85.2%, and a negative predictive value (NPV) of 48.1%. For comparison, when using the presence of IEDs on EEG (as per the EEG report) as the index test, the sensitivity is 37.0%, specificity is 100.0%, PPV is 100.0%, and NPV is 41.1%, with an AUROC of 0.69 (95% CI: 0.64–0.73). The AUROC of DeepEpilepsy was higher than any other method, although this was only statistically significant when compared to the ShallowConvNet models (AUROC: 0.60, 95%CI: 0.50–0.69). The diagnostic performances of all methods are shown in **Table 2**.

**Figure 3:**
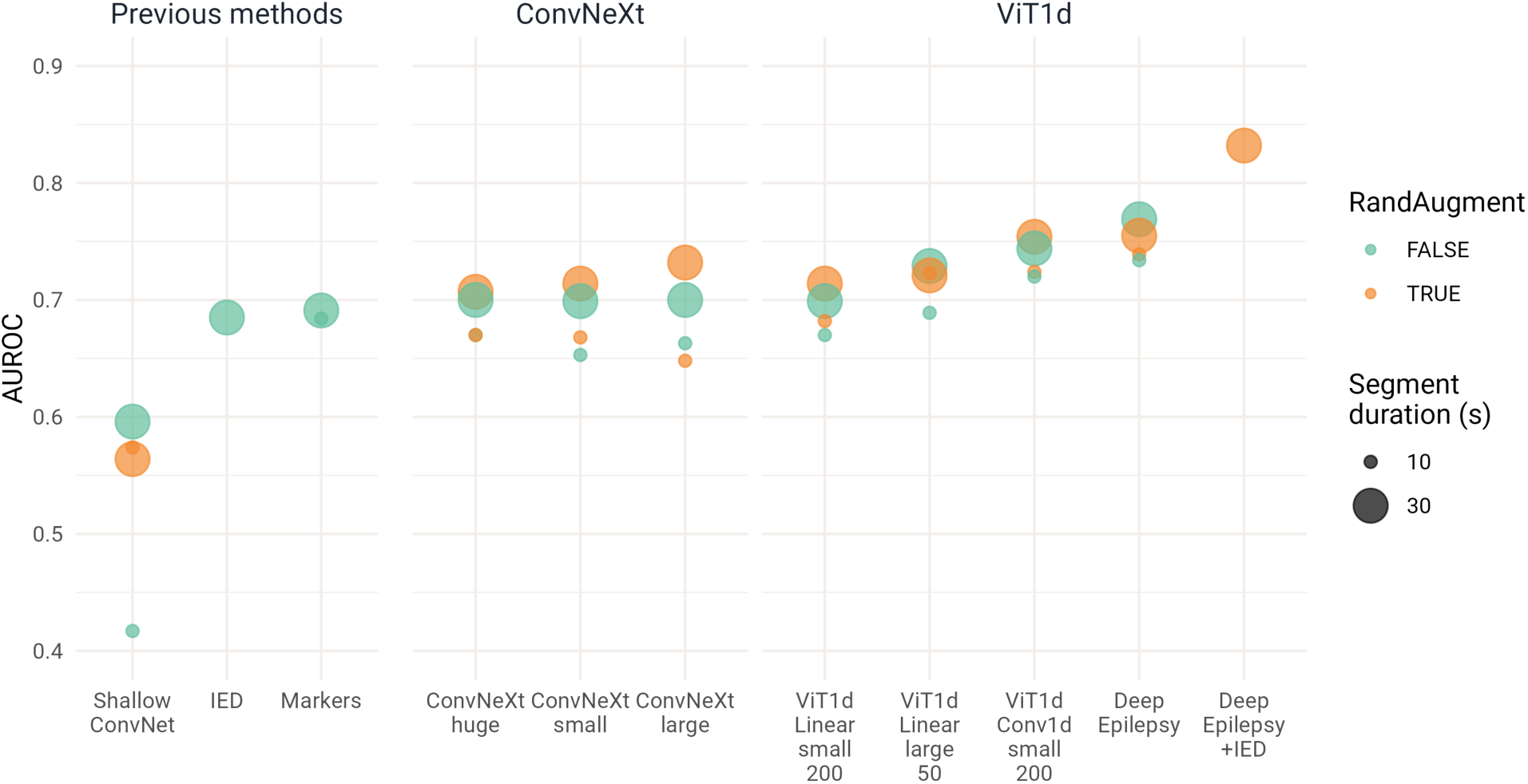
Diagnostic performances of automated EEG analysis for the diagnosis of epilepsy. Our flagship model, DeepEpilepsy, is shown alone and combined with traditional EEG interpretation based on the identification of IED. The other novel approaches shown are ViTs and ConvNeXt using different configurations (size: small, large, huge; tokenizers: convolutional or linear; window size: 50 pt or 200 pt) as well as presence of RandAugm and the duration of EEG segments used as input. Previous methods are the ShallowConvNet,^23^ extraction of computational markers,^21^ and the presence of IEDs on EEG. AUROC: Area under the receiver operating characteristic curve; IED: interictal epileptiform discharges; ViT: Vision Transformers.

**Table 1:** Description of the training (EEG recordings between January 2018 and July 2019) and testing cohorts (EEG recordings between July and September 2019)

|  | Training/validation cohort (n = 820) |  | Testing cohort (n = 128) |  |
| --- | --- | --- | --- | --- |
|  | Epilepsy | No Epilepsy | Epilepsy | No Epilepsy |
| Number of EEGs | 517 | 303 | 81 | 47 |
| Sex = woman (%) | 259 (50.1) | 159 (52.5) | 54 (66.7) | 26 (55.3) |
| Age (median [IQR]) | 42.00 [29.00, 58.00] | 57.00 [41.00, 67.00] | 37.00 [25.00, 57.00] | 60.00 [50.50, 71.00] |
| Total follow-up after EEG in weeks (median [IQR]) | 133.50 [95.75, 173.00] | 59.00 [17.00, 116.00] | 99.50 [70.25, 125.00] | 62.00 [17.00, 102.00] |
| Epilepsy type (%) |  |  |  |  |
| Focal | 370 (71.6) | — | 49 (60.5) | — |
| Generalized | 119 (23.0) | — | 26 (32.1) | — |
| Unknown | 28 (5.4) | — | 6 (7.4) | — |
| Age of epilepsy onset (median [IQR]) | 22.00 [13.00, 40.00] | — | 23.00 [14.00, 48.00] | — |
| Seizure recurrence after EEG (%) | 269 (52.0) | 0 (0.0) | 44 (54.3) | 0 (0.0) |
| Number of days since last seizure (median [IQR]) | 237 [56, 1134] | — | 118 [44, 467] | — |
| Number of epilepsy risk factors (median [IQR]) | 3 [1, 4] | 2 [1, 4] | 2 [1, 3] | 1 [0, 3] |
| History of epilepsy surgery (%) | 60 (11.6) | 0 (0.0) | 4 (4.9) | 0 (0.0) |
| Number of ASM (%) |  |  |  |  |
| 0 | 55 (10.6) | 253 (83.5) | 17 (21.0) | 42 (89.4) |
| 1 | 280 (54.2) | 36 (11.9) | 34 (42.0) | 5 (10.6) |
| 2 | 123 (23.8) | 12 (4.0) | 19 (23.5) | 0 (0.0) |
| 3 | 47 (9.1) | 2 (0.7) | 6 (7.4) | 0 (0.0) |
| 4 | 10 (1.9) | 0 (0.0) | 5 (6.2) | 0 (0.0) |
| 5 | 2 (0.4) | 0 (0.0) | 0 (0.0) | 0 (0.0) |
| Focal lesion on brain imaging (%) | 223 (43.1) | 84 (27.7) | 31 (38.3) | 10 (21.3) |
| Sleep deprived EEG (%) | 62 (12.0) | 50 (16.5) | 22 (27.2) | 8 (17.0) |
| IED (%) |  |  |  |  |
| Absence | 333 (64.4) | 282 (93.1) | 42 (51.9) | 46 (97.9) |
| Presence | 139 (26.9) | 2 (0.7) | 30 (37.0) | 0 (0.0) |
| Uncertain | 45 (8.7) | 19 (6.3) | 9 (11.1) | 1 (2.1) |
| Abnormal slowing on EEG (%) | 199 (38.5) | 46 (15.2) | 32 (39.5) | 10 (21.3) |

**Table 2:** Classification performances on the testing set for all machine learning methods.

|  | Segment duration (s) | RandAugment | AUC |
| --- | --- | --- | --- |
| **DeepEpilepsy** | 30 | False | 0.77 (0.69--0.84) |
| **DeepEpilepsy** | 30 | True | 0.76 (0.68--0.83) |
| ViT1d, Conv tokenizer, small | 30 | True | 0.75 (0.68--0.83) |
| ViT1d, Conv tokenizer, small | 30 | False | 0.74 (0.66--0.82) |
| **DeepEpilepsy** | 10 | True | 0.74 (0.66--0.81) |
| **DeepEpilepsy** | 10 | False | 0.73 (0.64--0.81) |
| ConvNeXt, large | 30 | True | 0.73 (0.65--0.81) |
| ViT1d, Linear tokenizer, large | 30 | False | 0.73 (0.65--0.80) |
| ViT1d, Conv tokenizer, small | 10 | True | 0.72 (0.64--0.80) |
| ViT1d, Linear tokenizer, large | 10 | True | 0.72 (0.64--0.80) |
| ViT1d, Linear tokenizer, large | 30 | True | 0.72 (0.64--0.80) |
| ViT1d, Conv tokenizer, small | 10 | False | 0.72 (0.64--0.80) |
| ConvNeXt, small | 30 | True | 0.71 (0.63--0.80) |
| ViT1d, Linear tokenizer, small | 30 | True | 0.71 (0.63--0.79) |
| ConvNeXt, huge | 30 | True | 0.71 (0.62--0.79) |
| ConvNeXt, huge | 30 | False | 0.70 (0.61--0.78) |
| ConvNeXt, large | 30 | False | 0.70 (0.62--0.78) |
| ViT1d, linear tokenizer, small | 30 | False | 0.70 (0.61--0.78) |
| ConvNeXt, small | 30 | False | 0.70 (0.61--0.78) |
| Feature extraction with LightGBM | 30 | --- | 0.69 (0.60--0.78) |
| ViT1d, Linear tokenizer, large | 10 | False | 0.69 (0.60--0.76) |
| Feature extraction with LightGBM | 10 | --- | 0.68 (0.59--0.77) |
| ViT1d, linear tokenizer, small | 10 | True | 0.68 (0.59--0.76) |
| ConvNeXt, huge | 10 | False | 0.67 (0.58--0.76) |
| ConvNeXt, huge | 10 | True | 0.67 (0.58--0.75) |
| ViT1d, linear tokenizer, small | 10 | False | 0.67 (0.58--0.75) |
| ConvNeXt, small | 10 | True | 0.67 (0.58--0.76) |
| ConvNeXt, large | 10 | False | 0.66 (0.58--0.75) |
| ConvNeXt, small | 10 | False | 0.65 (0.57--0.74) |
| ConvNeXt, large | 10 | True | 0.65 (0.56--0.73) |
| ShallowConvNet | 30 | False | 0.60 (0.49--0.69) |
| ShallowConvNet | 10 | True | 0.57 (0.47--0.67) |
| ShallowConvNet | 30 | True | 0.56 (0.46--0.66) |
| ShallowConvNet | 10 | False | 0.42 (0.32--0.51) |

When using the two-step model as the index test (1: presence of IED classified as epilepsy, 2: if no IED: DeepEpilepsy models prediction), the AUROC was 0.83 (95%CI: 0.77–0.89; **Figure 3**). The sensitivity, specificity, PPV, and NPV were 73.2%, 74.5%, 86.7%, and 55.1%.

### Subgroup analyses

In the subgroup of 77 patients not diagnosed with epilepsy at the time of the EEG, DeepEpilepsy still had above-chance performances (AUROC: 0.69, 95%CI 0.56–0.80), and the two-step model had the following performances: sensitivity of 65.6%, specificity of 76%, PPV of 63.6% and NPV of 77.6%, with an AUROC of 0.77 (0.65–0.87). The ROC curves for IEDs only, DeepEpilepsy, and DeepEpilepsy combined with IEDs for this subgroup are shown in **Figure 4**.

**Figure 4:**
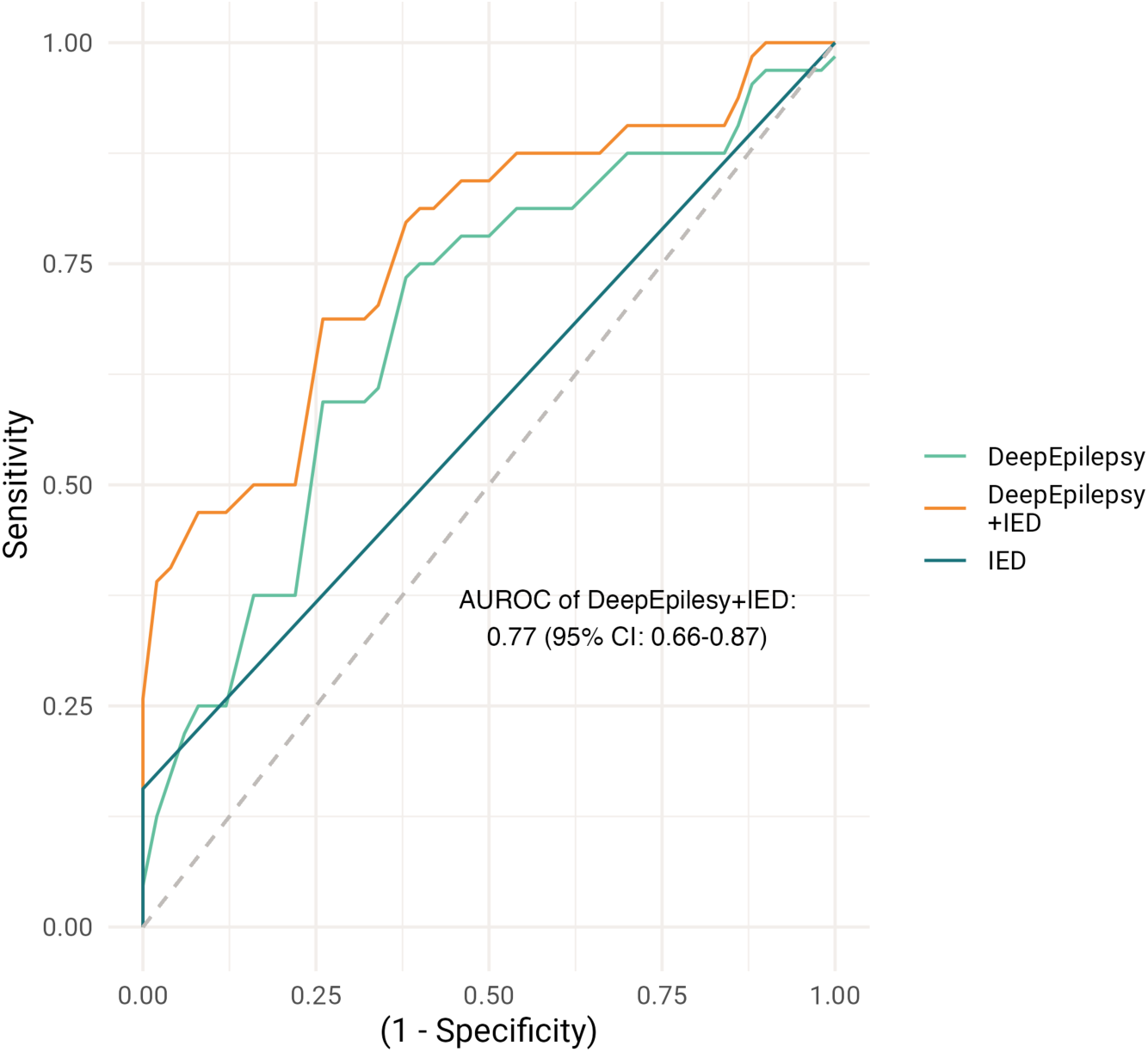
ROC curves for IEDs only, DeepEpilepsy, and DeepEpilepsy combined with IEDs in the subgroup of patients not diagnosed with epilepsy at the time of the EEG (*n* = 77). AUROC: Area under the receiver operating characteristic curve; IED: interictal epileptiform discharges.

The results for other subgroups are presented in **Figure 5**. Notably, in absence of IEDs, AUROC was 0.74 (0.65–0.83). Across other subgroups, performances were above chance except for patients > 60 years old and patients with a single antiseizure medication.

**Figure 5:**
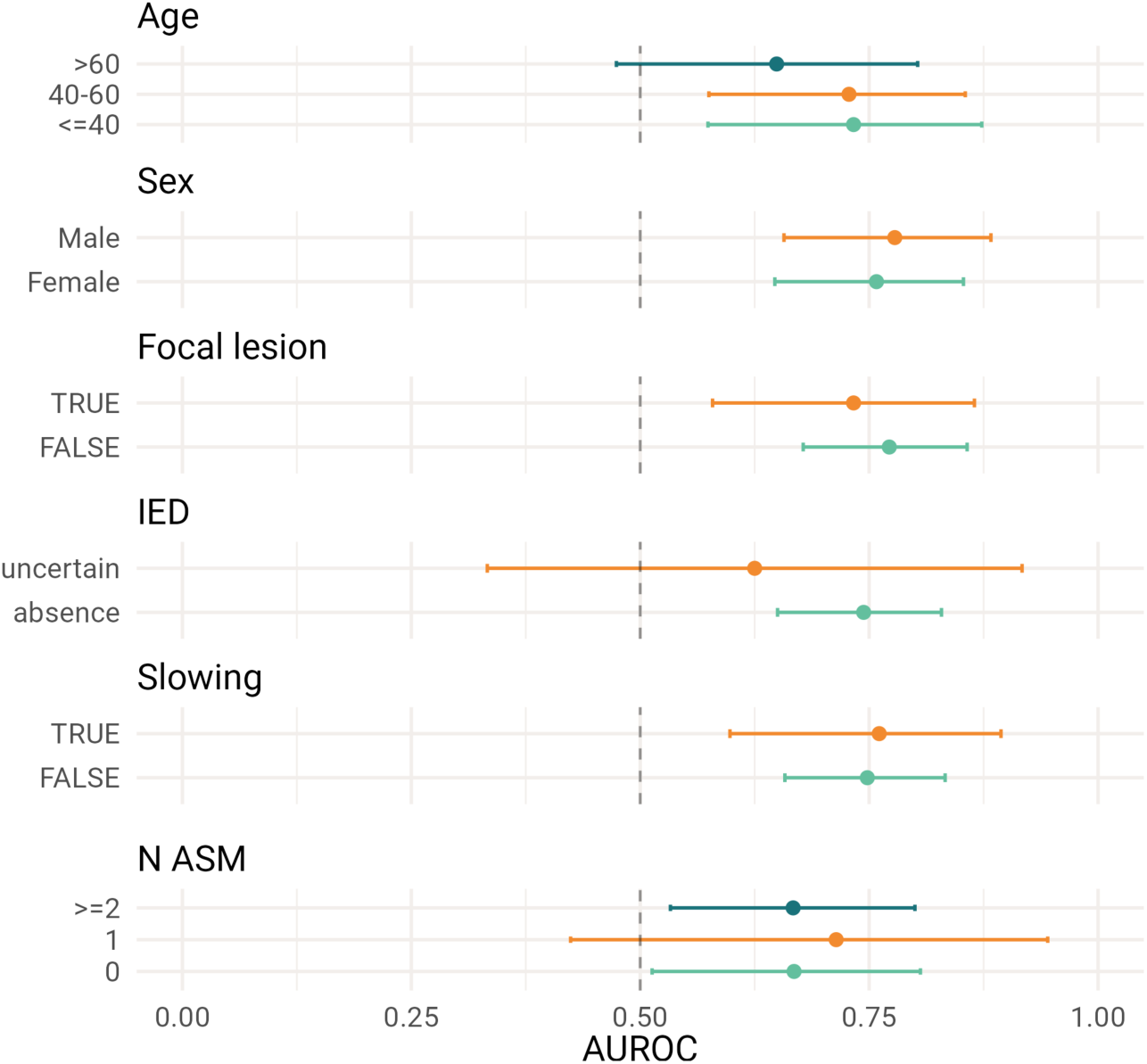
Performance of DeepEpilepsy for classification of epilepsy diagnosis from routine EEG in different subgroups of the testing set. ASM: Antiseizure medication; AUROC: Area under the receiver operating characteristic curve; IED: interictal epileptiform discharges.

### Sample size analysis

We trained the different neural network models on subsets of the data (50, 100, 250, 500, and 750 EEGs) to assess the impact of the size of the training sample on performance (**Figure 6**). With 10-second segments, the ShallowConvNet had highest performances when trained on 250 EEG recordings. The other models tended towards increased performances, with a ceiling at 500 EEGs. Using 30-second segments, the ShallowConvNet showed a slight increase in performances with increased training size, with a maximal AUROC of 0.6 at 750 EEGs. In contrast, the performance of the ConvNeXt and ViT models increased almost linearly with sample size, achieving the highest performances with 750 EEGs. In almost all cases, 500 EEGs was the minimal training size required to achieve above-chance performances.

**Figure 6:**
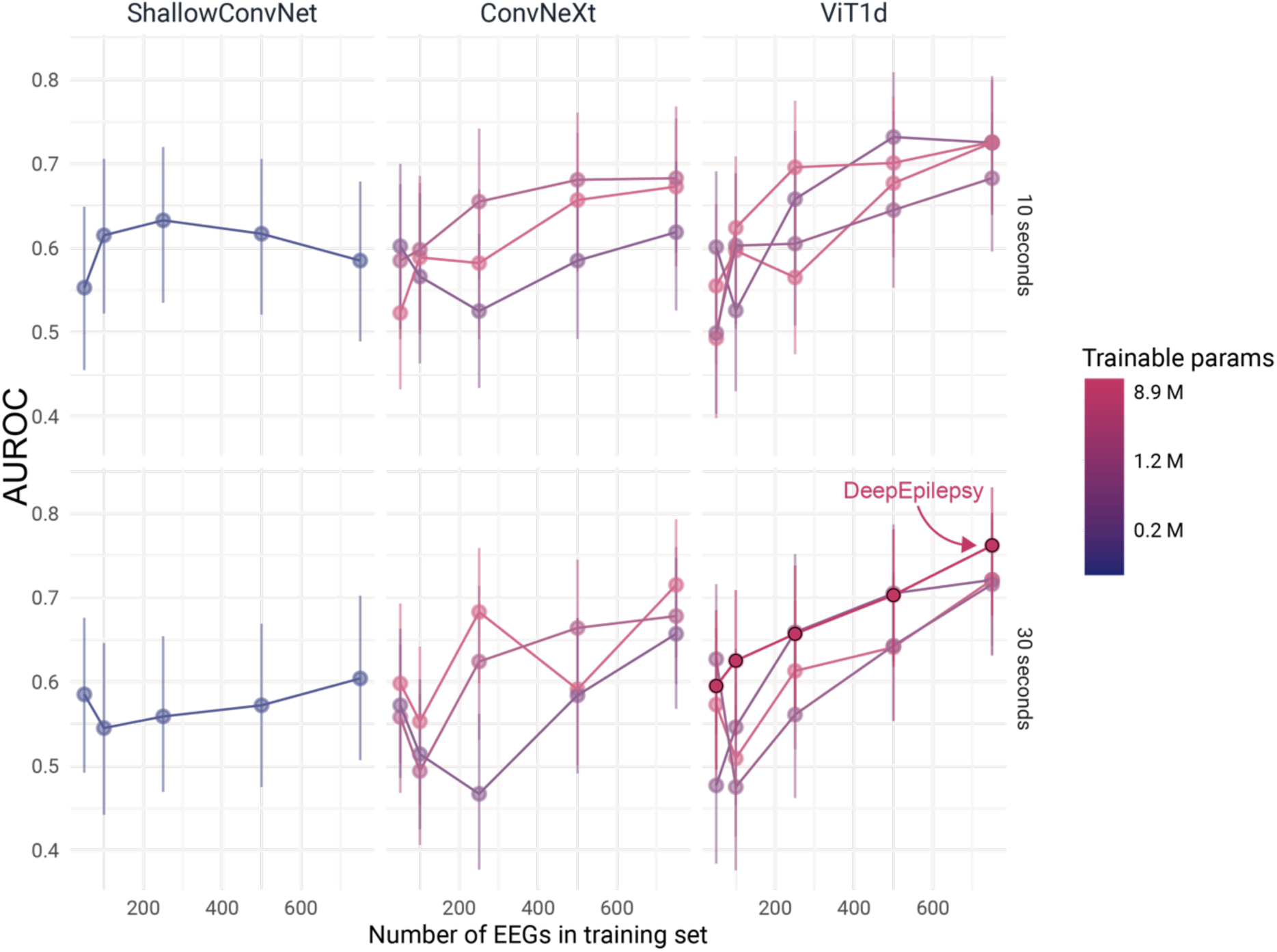
Impact of training sample size on the performance of four deep learning models (ShallowConvNet, ConvNeXt, ViT1d, and DeepEpilepsy) for detecting epilepsy from EEG segments. Performance is measured by the AUROC score, with models trained on varying numbers of EEGs (50, 100, 250, 500, and 750). The models were trained on 10s (top row) and 30s (bottom row) overlapping EEG segments. AUROC: Area under the receiver operating characteristic curve; IED: interictal epileptiform discharges; ViT: Vision Transformers.

For reference, using our segmentation strategy, 500 EEGs resulted in 765,000 10-second overlapping segments or 500,000 30-second overlapping segments.

### Relationship between learned representations and traditional EEG features

We analyzed the band power and entropy of EEG segments in relation to their distribution in latent space of Deep Epilepsy and ShallowConvNet using a clustering algorithm. For band power, DeepEpilepsy produced clusters with higher variance in the high-frequency range (> 13 Hz), particularly in the 20–40 Hz, 40–75 Hz, and 75–100 Hz bands. In contrast, ShallowConvNet exhibited relatively higher variance in the low-frequency range (< 10 Hz) (**eFigure 2**). Although DeepEpilepsy showed significant heterogeneity across all frequency bands, ShallowConvNet had non-significant analysis of variance in the 20–40 Hz range (*p* = 0.24) Regarding entropy, both models showed significant heterogeneity across all frequencies, but ShallowConvNet displayed higher inter-cluster variance, especially for bands above 1.6 Hz, suggesting that this was a key feature learned by this model (**eFigure 3**).

## Discussion

This study assessed the diagnostic performance of DL-based analysis of routine EEG for epilepsy. We developed and trained the DL models on 948 consecutive EEGs from 846 patients, testing them on a temporally shifted cohort of 128 EEGs from 118 patients. Our flagship model, DeepEpilepsy, had a testing AUROC of 0.76 (95%CI: 0.69–0.83), outperforming other methods including conventional IED- based interpretation and previously proposed computational methods. Combining the presence of IEDs with DL analysis increased the AUROC to 0.83 (95%CI: 0.77–0.89), demonstrating a potential for clinical translation.

Epilepsy diagnosis is primarily clinical, guided by individualized seizure recurrence risk assessment, which can be challenging due to limited reliable data.^1^ The identification of IEDs on rEEG is commonly used to support the diagnosis of epilepsy, but their low sensitivity and risk of over-interpretation can often lead to both over- or underdiagnosis.^11^ In our study, IEDs had an AUROC of 0.69 with a sensitivity as low as 37%. Our DL models provided higher overall diagnostic performances from the EEG than IEDs.

Combining both approaches allowed to leverage the model’s higher sensitivity and the high specificity of IEDs. Currently, no definitive, quantitative, non-ictal biomarkers have been validated for clinical use.^1^ Although several studies have explored changes in the EEG such as shifts in band power^15,26,27^ or changes in entropy,^28,29^ many remain at the “proof-of-concept” stage, limited by case-control designs or inadequate validation.^13^ More recent studies on computational analysis of EEG for the diagnosis of epilepsy have shown mixed results.^12,30^ Unlike prior work,^13^ our validation cohort corresponds to the group of patients in which the algorithm would be used in real-life, reducing bias in performance evaluation. Furthermore, the gold-standard in our study was based on a thorough review of clinical notes with a median follow-up period of over two years, allowing the clinician to build a more complete clinical picture integrating seizure recurrence, imaging, video-EEG evaluations, or new clinical symptoms. This is in contrast with studies that based the diagnosis on the EEG report or a single clinical visit.^13^ These methodological strengths reduce bias and represent key steps towards the clinical integration of automated EEG analysis.^13^

DeepEpilepsy is based on the Transformer architecture,^31^ which has greatly advanced our capacity to model sequence data. Transformers have been adapted for EEG-based tasks such as eye-tracking,^32^ seizure prediction,^33,34^ and decoding of motor patterns.^35^ A critical component in adapting Transformers to EEG is the tokenization method, which influences feature extraction and the timescales captured by the model. Previous studies have used separable convolutions as the tokenizer,^37,40^ a popular approach in EEG models since the ShallowConvNet and EEGNet CNNs.^17,41^ However, in our early experiments, we found this approach underperformed and was inefficient, leading us to discard it. In contrast to the original ViT model, which “patchified” the input signal with a linear, non-overlapping tokenizer,^25^ we showed that a deep convolutional embedding results in higher performances. This improvement is likely due to the convolution’s inductive bias towards hierarchical dynamics across timescales and spatial scales.^42^ The discrepancies between our findings and previous studies on Transformer-based EEG models probably arise, in part, from dataset size and complexity: our training dataset included over 1 million samples from more than 900 patients, while prior studies used significantly smaller training samples (15 000–80 000 segments from 23–70 patients^37–39,43^) as well as shorter EEG segments (up to 50 000 points,^37–39,43^ compared to our 114,000 points per segment).

A notable advantage of Transformers over CNNs is their scalability. DeepEpilepsy showed continual improvement as the size of the training sample increased, without hitting a performance ceiling. Recent studies have further demonstrated CNNs’ limitations in scaling to large EEG datasets.^44^ The absence of a performance ceiling in DeepEpilepsy suggests potential for further improvements with larger datasets, motivating multicenter collaborations to expand the training sample.

Unlike other approaches to automated EEG interpretation,^19,20^ DeepEpilepsy did not rely on IEDs. We hypothesize that DeepEpilepsy’s capacity to detect epilepsy may be linked to changes in the higher frequency spectrum (40–100 Hz). The gamma range hosts high-frequency oscillations (HFOs). While they may have a prognostic value in patients with refractory temporal lobe epilepsies,^37,38^ HFOs are not captured by our frequency range. All models performed better with longer EEG segments (30s), a timescale typically outside of the scope of routine EEG interpretation, suggesting that the models may capture brain dynamics not traditionally considered and warranting further investigations.

Integrating DL models like DeepEpilepsy in the clinical workflow could enhance clinical decision-making by the increasing the information available in case of diagnostic uncertainty. For example, a positive prediction by the model in a patient with neurological events of uncertain significance and negative workup (no IEDs on EEG, no epileptogenic lesion on MRI) could increase the suspicion of epilepsy, prompting to more frequent follow-ups or repeat EEGs. Conversely, a patient with a low pre-test probability of epilepsy, absence of IEDs and a negative DL prediction could reduce clinical suspicion.

Most likely, combined with advances in other domains such as text processing, imaging and genetics,^44–46^ the automated EEG analysis will lead to a more comprehensive phenotyping of these patients and potentially lead to quantifying the seizure likelihood. This could also improve clinical trials in epilepsy, which are currently limited by self-reported and unreliable outcome measures.^4,47^

This study has limitations. Our data comes from a single center, and although routine EEG recording is standardized, variability in hardware, software, and technique may affect generalizability. Additionally, at our center, patients with a first unprovoked seizure presenting at the emergency department generally undergo their EEG there and not as outpatient, limiting their inclusion in our cohort. Another limitation is the use of the EEG report as a measure of whether an EEG contains IEDs, which could be biased as EEG readers are not blinded to the diagnosis. However, for patients which were “undiagnosed” at the time of the EEG, the limitation does not apply. Finally, subgroup analyses were limited by the relatively small sample size.

In conclusion, this study demonstrates that DeepEpilepsy, a Transformer model, could identify epilepsy on routine EEG independently of IEDs. The DL algorithm alone had an AUROC of 0.76, surpassing previously proposed methods, which was increased to 0.83 when combined with IEDs. Several questions remain such as the exact nature of brain dynamics captured by DeepEpilepsy, the optimal sample sizes for training the model, and the true clinical impact of this increased diagnostic yield in specific clinical settings.

## Supporting information

STARD Checklist

## Acknowledgements

We would like to acknowledge the work of the CHUM EEG technologists for their contribution to the recording of the EEGs. We would also like to thank Manon Robert and Véronique Cloutier for their help regarding the access to the EEG data and the submission to the ethics review board.

## Author contributions

EL, DT, DKN, FL, anf EBA conceived and planned the experiments. EL, AQX, MJ, and JDT collected the data. EL, AQX, MJ, JDT, and EBA had direct access and verified the underlying data. EL performed the experiments. EL, DT, DKN, FL, and EBA contributed to the interpretation of the results. EL wrote the first draft of the manuscript. All authors provided critical feedback and reviewed the manuscript.

