## Supplementary Material for "Improving Diagnostic Accuracy of Routine EEG for Epilepsy using Deep Learning"

#### eMethod 1: Automated processing of EEG and classification

**Deep Learning:** We implemented two DL approaches. First, we adapted to EEG data the ConvNeXt model, a deep CNN analog to the ResNet, selected for its robust performance in computer vision tasks.[1] Second, we implemented a novel model coined *DeepEpilepsy*, based on the Vision Transformer (ViT) architecture, a Transformer model that takes images as input and that outputs class probabilities.[2] DeepEpilepsy uses a three-layer convolutional tokenizer plus a bottleneck convolution, restricting the complexity of the model and allowing to capture multi-scale features, akin to the Compact Convolutional Transformer.[3] We also tested a tokenizer with non-overlapping linear patch embedding proper to the original ViT model.[2]

For all DL models, EEGs were segmented into overlapping 10- or 30-second segments and scaled so that each channel had a mean of zero and standard error of one. These scaled segments were used as input into the DL models. To enhance model generalization, we applied a random data augmentation algorithm during training, similar to the RandAugm algorithm.[4] For each EEG segment, one augmentation was drawn randomly from a set of transformations, which included filtering (band-pass, low-pass, high-pass), masking (channel, time), and adding noise (**eFigure 1**). These augmentations were applied with a 50% probability. The intensity of the augmentation (e.g., filter frequency, noise level, mask length) was also randomized and controlled by a hyperparameter M. Based on initial experiments on the training and validation data, we set M = 8.

We performed a Bayesian hyperparameter search on the training and validation set to select four different configurations for the ViT (one of which was DeepEpilepsy) (**eTable 1**) and three for ConvNeXt (**eTable 2**). We also investigated different learning rates, weight decay, and batch size values. The final models were trained on the entire training and validation set. The optimization hyperparameters and model specifications are described in **eTable 4**.

**ShallowConvNet:** We reimplemented the ShallowConvNet model following the configuration outlined in Schirrmeister et al.[5] However, after conducting a hyperparameter search on the training and validation set, we identified a more optimal configuration specific to our dataset, which we used for testing (**eTable 3**). The EEG segmentation and standardization were consistent with the other DL models. Similarly, we optimized training hyperparameters (learning rates, weight decay, and batch size values) through a Bayesian search on the training and validation set (**eTable 4**).

**EEG markers:** We followed the methodology described in Lemoine et al.,[6] selecting only the best-performing markers and testing both 10- and 30-second segments. EEGs were segmented at pre-specified time points (every change of montage, every 15s during hyperventilation, every 15s for two minutes post-hyperventilation, every photic stimulation frequency, and every eye closure or opening). We applied an automated artifact detection/rejection algorithm (*AutoReject*)[7] and extracted the following markers: fuzzy entropy, line length, correlation dimension, band power, and peak alpha. Band power was calculated using a multitaper method, with integrals estimated using Simpson's method (frequency ranges: 100–75 Hz, 75–40 Hz, 40–20 Hz, 20–13 Hz, 13–10 Hz, 10–8 Hz, 8–6 Hz, 6–4 Hz, 4–2 Hz, and 2–1 Hz). For nonlinear features (fuzzy entropy, line length, and correlation dimension), the *Sym5* wavelet was used with six decomposition levels (with frequency ranges: 100–50 Hz, 50–25 Hz, 25–12.5 Hz, 12.5–6.25 Hz, 6.25–3.125 Hz, and 3.125–1.56 Hz).[8] One value was extracted per marker, EEG, segment, channel, and frequency band. Missing values were imputed using multivariate iterative imputation. Markers were used as input features for an L1-regularized boosted-trees classifier (LightGBM). The optimal hyperparameters for the classifier were selected via Bayesian optimisation using a 5-fold cross-validation on the training and validation set.

#### eMethod 2: Interpretability

We performed an exploratory analysis of the embeddings learned by DeepEpilepsy to better understand which patterns were captured by the DL model. Thirty-second segments from the testing set were processed through DeepEpilepsy, and their embeddings were extracted. A clustering algorithm was then used to group the embeddings into 12 distinct clusters. We computed the band power and entropy of the input segments and compared the distribution of values between clusters. To compute band power and entropy, we used the same methods and frequency ranges as in the previous section **Automated processing of EEG and classification: EEG markers**. We repeated this procedure with the ShallowConvNet model. To test for heterogeneity between clusters, we applied an analysis of variance (Krusper-Wallis test) at each frequency bands. We then compared the F-score between both models and between frequency bands.

#### eFigure 1: Data augmentations used by the RandAugm algorithm

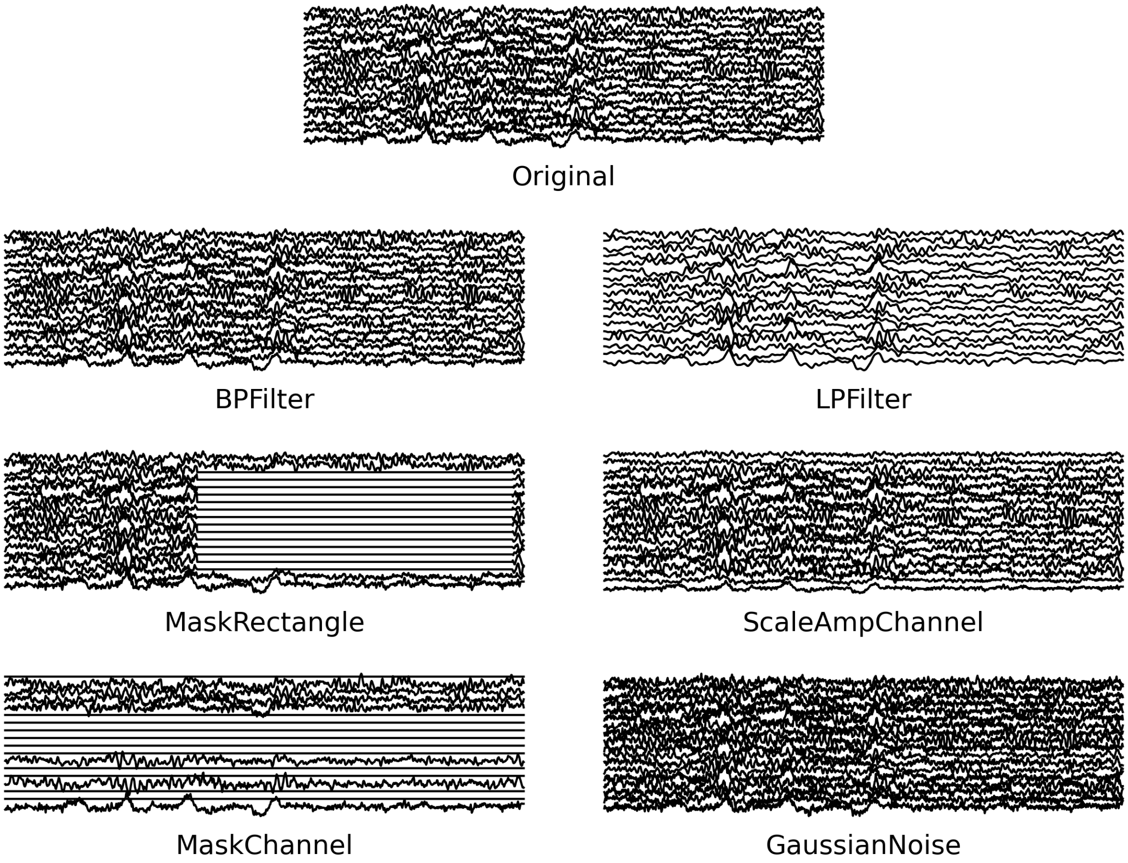


**eFigure 1:** Data augmentations used by the RandAugm algorithm, alongside the original EEG sample. BPFilter: band pass filter with randomly chosen frequency window. LPFiler: Low-pass filter, with random cut-off frequency. MaskRectangle: masking of data points contiguous in both time and space. ScaleAmpChannel: Random scaling of channels. MaskChannel: masking of all data points in randomly selected channels. GaussianNoise: Addition of gaussian noise with a random intensity. The intensity of the augmentations is scaled according to a hyperparameter M. For example, higher values of M result, on average, in lower values of cutoff frequencies for LPFilter, larger mask area for MaskRectangle, and a larger number of channels affected by ScaleAmpChannels as well as a higher amplitude of scaling.

### eTable 1–4: Deep learning hyperparameters for the final model configurations

**eTable 1:** Model configurations for the Vision Transformer (ViT) models

| Model | patch size | tokenizer | tokenizer: layers | hidden dim | layers | heads | MLP size | dropout | attention dropout | params (M) |
|---|---|---|---|---|---|---|---|---|---|---|
| ViT1d, linear, small | 200 | Linear | 1 | 128 | 2 | 2 | 128 | 0.25 | 0.25 | 0.7 |
| ViT1d, linear, large | 50 | Linear | 1 | 512 | 6 | 8 | 512 | 0.25 | 0.25 | 10.0 |
| ViT1d, Conv, small | 200 | Convolution | 3 | 128 | 2 | 2 | 128 | 0.25 | 0.25 | 0.4 |
| DeepEpilepsy: ViT1d, Conv, large | 50 | Convolution | 3 | 512 | 6 | 8 | 512 | 0.25 | 0.25 | 10.6 |

**eTable 2:** Model configurations for the ConvNext models

| Model | blocks | channels | stem: downsampling scale | drop path rate | params (M) |
|---|---|---|---|---|---|
| ConvNeXt, small | 1, 1, 3, 1 | 16, 32, 64, 128 | 4 | 0.1 | 0.3 |
| ConvNeXt, large | 2, 2, 6, 2 | 32, 64, 128, 256 | 2 | 0.1 | 2.0 |
| ConvNeXt, huge | 3, 3, 9, 3 | 64, 128, 256, 512 | 2 | 0.1 | 11.9 |

**eTable 3:** Model configuration for the ShallowConvNet model

| Model | kernel size (stride) | space conv channels | time conv channels | max pool window | dropout | params (M) |
|---|---|---|---|---|---|---|
| ShallowConvNet | 16 (1) | 64 | 128 | 80 | 0.25 | 0.040 |

**eTable 4:** Optimization parameters for all neural networks

| Parameter | value |
|---|---|
| Optimizer | AdamW |
| Base learning rate | 1.0e-5 |
| Weight decay | 0.05 |
| Optimizer momentum | β1, β2=0.9, 0.999 |
| Batch size | 512 |
| Training epochs | 30 |
| Learning rate schedule | Cosine decay |
| Warmup iterations | 1000 |
| Warmup schedule | Linear |
| RandAugm M | 7 |
| Gradient clipping | 1.0 (ViT only) |

### eTable 5: Clinical characteristics of the "undiagnosed" subgroup of the testing cohort

| | Epilepsy | No Epilepsy |
|---|---|---|
| Number of patients | 28 | 47 |
| Sex = woman (%) | 19 (67.9) | 26 (55.3) |
| Age (median [IQR]) | 41.00 [34.75, 58.25] | 60.00 [50.50, 71.00] |
| Total follow-up after EEG in weeks (median [IQR]) | 119.00 [95.00, 134.50] | 62.00 [17.00, 102.00] |
| Epilepsy type (%) | | |
| Focal | 23 (82.1) | – |
| Generalized | 3 (10.7) | – |
| Unknown | 2 (7.1) | – |
| Age of epilepsy onset (median [IQR]) | 37.00 [23.25, 52.00] | – |
| Seizure recurrence after EEG (%) | 17 (60.7) | – |
| Number of days since last seizure (median [IQR]) | 87.50 [33.00, 164.00] | – |
| Number of epilepsy risk factors (median [IQR]) | 2.00 [1.00, 4.00] | 1.00 [0.00, 3.00] |
| History of epilepsy surgery (%) | 0 (0) | – |
| Number of ASM (%) | | |
| 0 | 9 (32.1) | 42 (89.4) |
| 1 | 12 (42.9) | 5 (10.6) |
| 2 | 5 (17.9) | 0 (0.0) |
| 3 | 2 (7.1) | 0 (0.0) |
| 4 | 0 (0.0) | 0 (0.0) |
| 5 | 0 (0.0) | 0 (0.0) |
| Focal lesion on brain imaging (%) | 10 (35.7) | 10 (21.3) |
| Sleep deprived EEG (%) | 9 (32.1) | 8 (17.0) |
| IED (%) | | |
| Absence | 12 (42.9) | 46 (97.9) |
| Presence | 10 (35.7) | 0 (0.0) |
| Uncertain | 6 (21.4) | 1 (2.1) |
| Abnormal slowing on EEG (%) | 10 (35.7) | 10 (21.3) |

#### eFigure 2: Power spectrum density of EEG segments clustered according to their latent representations using DeepEpilepsy vs. ShallowConvNet

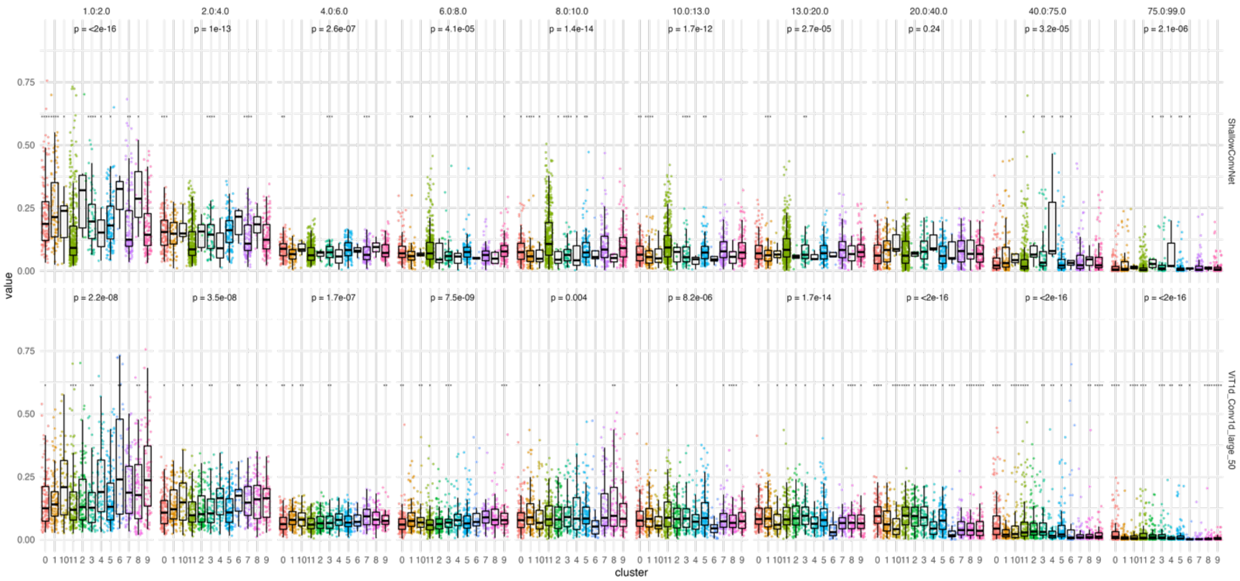


**eFigure 2:** Power spectrum density of 30s EEG segments clustered according to their latent representations using DeepEpilepsy vs. ShallowConvNet. Each EEG segments was processed through either the trained ShallowConvNet (top) or the trained DeepEpilepsy (ViT1d_Conv1d_large_50, bottom) to generate a latent vector. The latent vectors were then clustered using K-means clustering (K=12). The power in each band was calculated for the input segment (1 Hz:2 Hz, 2 Hz:4 Hz, etc.) and plotted on the y-axis. A statistical analysis of inter-cluster variance was perform in each frequency band using the Krusper-Wallis test (*p*-values at the top of each facet). A lower p-value correspond to a larger heterogeneity between clusters in that frequency bands.

#### eFigure 3: Entropy of EEG segments clustered according to their latent representations using DeepEpilepsy vs. ShallowConvNet

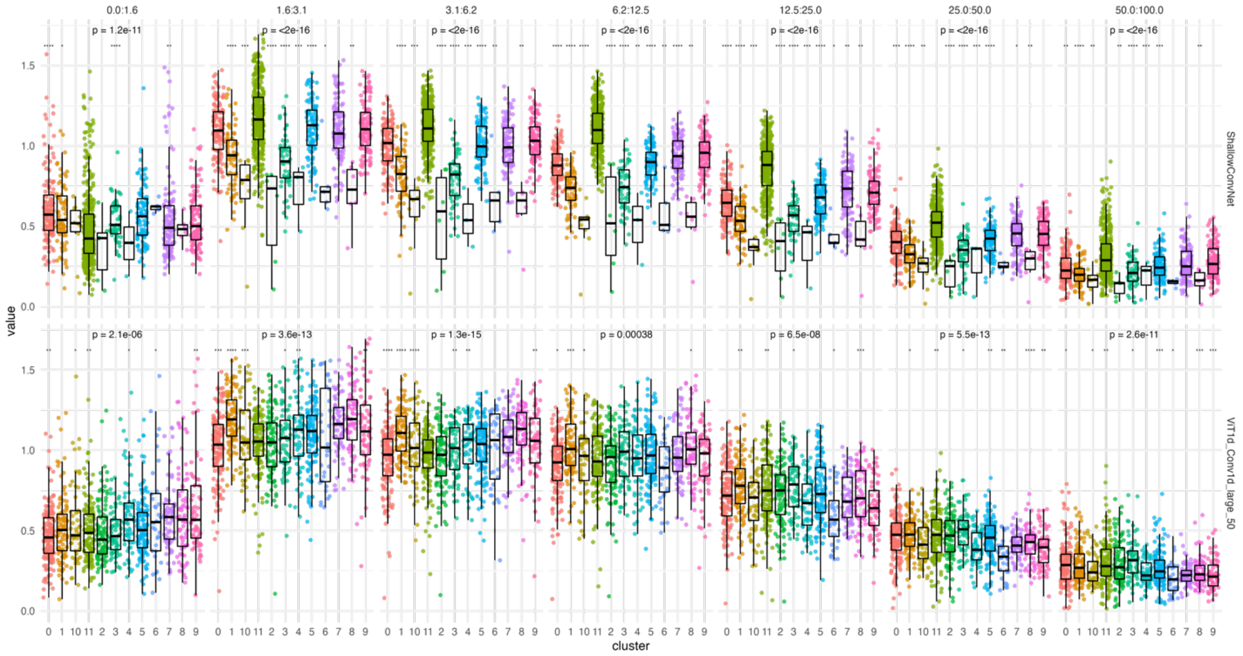


**eFigure 3:** Entropy of EEG segments clustered according to their latent representations using DeepEpilepsy vs. ShallowConvNet. Each EEG segments was processed through either the trained ShallowConvNet (top) or the trained DeepEpilepsy (ViT1d_Conv1d_large_50, bottom) to generate a latent vector. The latent vectors were then clustered using K-means clustering (K=12). The entropy in each band was calculated for the input segment (1 Hz:2 Hz, 2 Hz:4 Hz, etc.) and plotted on the y-axis. The fuzzy entropy algorithm was used with parameters *m*=2 and *r*=0.2. A statistical analysis of inter-cluster variance was perform in each frequency band using the Krusper-Wallis test (*p*-values at the top of each facet). A lower p-value correspond to a larger heterogeneity between clusters in that frequency bands.
